# Frequency of antimicrobial resistance among fecal bacteria pathogens in an informal settlement in Nairobi, Kenya, 2018 to 2020

**DOI:** 10.1101/2025.06.01.25328746

**Authors:** Bonventure Juma, Matthew Mikoleit, Newton Wamola, Mike Powel Osita, Caroline Ochieng, Samuel Kariuki, Elizabeth Hunsperger

## Abstract

**Background:** Multidrug-resistant bacterial enteric pathogens such as *Salmonella* and *Shigella* have the potential to cause significant mortality and represent a major issue facing the global health community. This is particularly concerning in low-and middle-income countries where access to clean water and antimicrobials is limited. We aimed to determine the frequency, antimicrobial resistance, and presence of resistance genes among *Salmonella, Shigella, Vibrio, & Campylobacter* recovered from patients presenting with diarrhea in an informal settlement in Nairobi, Kenya, from 2018 to 2020.

**Methodology:** Conventional bacteriologic methods were used for bacterial culture and isolation. BD Phoenix M50 technology was used for the identification and speciation of bacteria recovered from stool samples and for determining minimum inhibitory concentrations to both clinically relevant antimicrobials and agents of epidemiologic significance. PCR testing was subsequently performed to identify resistance genes.

**Results:** The key pathogenic bacteria recovered were: *Shigella flexneri (58*.*3%), Salmonella enterica* serovar Typhi (8.3%), *Shigella dysenteriae (6*.*7%), Shigella boydii (6*.*7%)*, and *Shigella sonnei*, (6.7%); no *Campylobacter* or *Vibrio* were isolated. Overall resistance among *Shigella spp*. and *Salmonella spp*. was determined to be highest to ampicillin (83%), followed by tetracycline (50%), cotrimoxazole (47%), chloramphenicol (17%), ceftriaxone (7%), gentamycin (5%), meropenem (2%), ciprofloxacin (0%) and amoxicillin-clavulanic acid (2.0%). Resistance genes detected included β-lactamases (*bla*_TEM_ and *bla*_SHV_), aminoglycoside 3-N acetyltransferase *acc*(3), sulfonamide resistance genes (*sul*1, *sul*2), and tetracycline resistance genes *tet*(A) and *tet*(B).

**Conclusion:** Results emphasize the critical need for active and continuous surveillance of pathogenic enteric bacteria to improve patient management and inform the development of empiric therapy guidelines.

## Introduction

Antimicrobial resistance (AMR) has become a significant public health threat. Multidrug-resistant infections significantly impact patient management [1]. Treatment failure is particularly common in low- and middle-income countries (LMIC), potentially resulting in increased morbidity and mortality [2,3]. The threat is greater in LMICs because of the higher burden of bacterial illness, delayed health-seeking behavior, delayed access to laboratory diagnostics, unregulated access to antibiotics without a prescription, failure to comply with drug dosage/duration guidelines, high cost, and reduced availability of second-line antibiotics [4].

One critical aspect of the global response to AMR is surveillance. According to a 2017 World Health Organization-Africa region report, Africa has one of the largest data gaps on the prevalence of AMR [5,6]. This is attributed to a lack of funds to carry out surveillance, insufficient surveillance networks and inconsistent strategies to conduct AMR surveillance, poor laboratory infrastructure for detection of AMR including scarcity of experienced workforce, limited quality assurance and control for antibiotic susceptibility tests (AST) for AMR, lack of resources to procure laboratory reagents, and lack of equipment [7]. Antimicrobial-resistant bacteria burden healthcare systems, leading to complications in patient management of infectious diseases. In Kenya and Mozambique, the incidence of multidrug resistance among Enterobacterales and non-fermenting gram negatives has been increasing over time, particularly to first-line antibiotics, aminoglycosides, cephalosporins, and quinolones [8-10].

Antimicrobial resistance genes can spread rapidly among different Enterobacterales as observed through horizontal transmission when different bacteria actively donate resistance traits to other organisms they interact with within the same environment [11-18]. Acquisition of these genes by pathogenic bacteria further limits the treatment options [19]. The transmission of resistance genes can happen in hospitals via nosocomial spread or in the community, particularly in those with poor water, sanitation, and hygiene conditions [10].

Enterobacterales are known to exhibit naturally occurring plasmid-mediated or chromosomal-mediated resistance mechanisms such as beta-lactamases, mutations leading to amino acid switches, or active efflux of the antimicrobial from the bacterial cell [15,20,21]. These resistance traits may be transferred and finally reach most of the Enterobacterales through a successful combination of mutagenesis with the capacity for antimicrobial resistance gene transfer as integrons, transposons, gene cassettes, or interactive conjugative elements [22,23]. Therefore, this threat calls for active AMR surveillance to detect and control the spread of resistance among pathogenic Enterobacterales. In Enterobacterales, these resistance genes are often contained on a mobile plasmid and cluster together on a large multi-resistant genetic region [16]. Resistant bacteria may carry a resistance mechanism, which is genetically expressed but may initially be phenotypically suppressed. This poses a problem in the clinical management of diseases caused by Enterobacterales as the patient may be treated with an ineffective antibiotic that will not manage the disease.

Studies in Kenya have documented multidrug-resistant Enterobacterales, but few have characterized the different resistance genes detected among these bacteria [8,10,24,25]. This study aimed to determine the frequency, antimicrobial resistance, and presence of resistance genes among *Salmonella, Shigella, Vibrio, & Campylobacter* recovered from patients presenting with diarrhea in an informal settlement in Nairobi, Kenya, from 2018-2020. The results will help inform on the need for enhanced detection so that these resistant organisms are identified, and infection prevention control measures and appropriate clinical management can be undertaken.

## Materials and methods

### Study design and site

This was a cross-sectional study of *Shigella, Salmonella, Vibrio*, and *Campylobacter* in the Kawangware informal settlement in Nairobi from Nov 2018 to Jan 2020. Kawangware is an under-resourced informal settlement situated about 15 kilometers northwest of Nairobi city between Lavington and Dagoretti. It is one of the largest informal urban settlements in Nairobi City, with a population of around 133,000 from diverse ethnic backgrounds (https://vision2030.go.ke/publication/third-medium-term-plan-2018-2022/). It is estimated that 65% of the population are children and youths. Most inhabitants live on less than the equivalent of 2 US dollars each day, and unemployment is high; many inhabitants are self-employed traders. The informal settlement is made up of improvised temporary dwellings often made from corrugated metal sheets and plywood, covering an area of 3 square kilometers. In addition to experiencing poverty, several factors associated with informal settlements, including overcrowding, substandard housing, poor water, sanitation, and hygiene, contribute to the increased incidence of infectious diseases and mortality among this population.

### Patient recruitment and eligibility

Patients were recruited from November 2018 to January 2020 in the satellite sub-county hospital serving Kawangware. Eligible patients for study inclusion were those with three or more episodes of loose, watery, mucoid, or bloody diarrhea in a day and who gave written consent to participate. After meeting the inclusion criteria, patients consented to study participation in a private room in the hospital. The informed consent process consisted of an explanation of the study by a nurse, including the risks and benefits of participating. Refusal to participate in the study did not deny them any benefits. Those who agreed to be included in the study were given a stool cup, given instructions on how to self-collect a stool sample, and directed to the private washroom in the facility for collection. For children under 18 years of age, written informed assent was provided by either their parent or guardian. Those who were unable to provide the stool at that time were given the stool cup to take home and instructions on how to collect the stool. As patients resided near the hospital, they were instructed to return to the hospital immediately after collection. If patients were unable to produce a stool specimen on the day of enrollment, they were instructed to collect the first stool the following morning and deliver it to the hospital immediately.

### Data collection

Study staff administered a detailed questionnaire to individual cases at the study site on demographics and risk factors for diarrhea in the study area. Completed study questionnaires were stored in a lockable cabinet, and the electronic data was secured in a password-encrypted laptop. Data was only accessible by the study’s Data Manager and Principal Investigator.

### Sample collection

The stool was collected in a sterile stool cup and care was taken to avoid contamination with urine, soil, or water. A portion of the collected stool (one full spatula) was transferred into the Cary-Blair transport medium and shipped immediately to the U.S. Centers for Disease Control and Prevention Kenya microbiology laboratory (Kibera-Nairobi) for processing and identification. The remaining stool was discarded and not stored. Inoculated Cary-Blair medium was discarded after plating the swab on primary media.

### Bacteria isolation and identification

Upon arrival at the laboratory, the stool samples were examined for firmness(loose/formed) and plated immediately on MacConkey (MAC), Xylose lysine desoxycholate (XLD), Thiosulfate bile esculin salt agar (TCBS), and Campylobacter blood agar and incubated for 18 to 24 hours at 37°C. Colonies morphologically consistent with *Salmonella* or *Shigella* were sub-cultured for purity on nutrient agar and then used for making the solution for the BD Phoenix M50 preparation for identification, minimum inhibitory concentration, and resistance gene detection. No *Campylobacter*-like or *Vibrio*-like colonies were observed.

### BD Phoenix M50 testing and antimicrobial susceptibility testing

Following purification from primary media, identification, and AST was performed using the BD Phoenix (BD Diagnostics, Sparks, MD) according to the manufacturer’s instructions. Pure colonies were suspended in 4.5mL Phoenix ID broth, vortexed, optical density read using BD Phoenix Spec Nephelometer set to a concentration between 0.5 to 0.6 McFarland and inoculated onto a BD Phoenix Gram-negative card.

A total of 30 antimicrobials were tested, but for this study, 11 were selected based on Clinical and Laboratory Standards Institute (CLSI) guidelines for the treatment of Gram-negative bacteria [26]. The breakpoints and interpretations were set based on the CLSI guidelines and MIC ranges of resistance recorded. *E. coli* ATCC 25922 was used as quality control in every run. The strains that were resistant to more than three categories of antimicrobials were designated as multidrug-resistant (MDR).

### Antimicrobial resistance gene testing

Bacterial isolates displaying phenotypic resistance were selected for molecular testing to detect antimicrobial resistance genes (ARG) conferring resistance to aminoglycosides, beta-lactams, fluoroquinolones, sulfonamides, and tetracyclines. PCR for ARGs was performed and analyzed as described by Van *et al* [27]. Briefly, amplifications were performed in a 25 μL reaction mix containing nuclease-free water, DreamTaq Green PCR master mix (CW Biotech, China), forward and reverse primers for sulfonamides, tetracyclines, beta-lactams and aminoglycosides ARGs were added to a final concentration of 0.5 μM. The DNA template was obtained from fresh bacterial suspensions cultured overnight.

### Statistical analysis

Descriptive statistics were presented as counts and percentages. The presence of each resistance phenotype and detection of associated resistance genes were presented as percentages.

### Communication of patient results

AMR results for documented pathogens (*Salmonella spp. & Shigella spp*.) were communicated back to the hospital through the clinician attached to the project. These were then availed to individual patients and utilized by the hospital to guide clinical management.

### Ethical consideration

Ethical approval for the study was granted by the Kenya Medical Research Institute – scientific and ethical review unit (Ref. No. KEMRI/SERU/CMR/079/3596), Centers for Disease Control and Prevention (#7163), and Washington State University institute review boards relying on the non-CDC IRB.

## Results

### Demographics of study subjects

A total of 1,209 patients were enrolled in the study. Among these (55.3%) were female, (40.3%) were male and 4.4% did not indicate. Tap water from a piped system into the compound was used for drinking by (59.6%) of respondents, (25.7%) reported using water from unprotected boreholes, (3.6%) reported drinking water directly from rivers and (4.4%) did not respond. For water treatment methods, (32.4%) reported boiling water before use, (26.9%) added chlorine and (4.9%) strained drinking water through a cloth or filter before use while 31% did not treat water before use. On the kind of toilet facility used, most study subjects reported using dry pit latrines with slab (52.3%) flush to to-pit latrines (39.7%), while fewer reported using ventilated improved toilets (2.8%) and open defecation (1.5%). When asked about their hand-washing practices after using the toilet, 82.4% of participants reported always washing their hands using water in buckets, while 1.1% responded that they sometimes washed their hands and 10.8% said they did not wash their hands. Most participants had completed some level of formal education: 87.3% had completed classes 1-4; 22.5% completed classes 5-8; 31.4% completed high school, and 5.5% had a tertiary education.

### Bacteria isolated

The most common pathogens identified were *Shigella flexneri* (2.7%), *Shigella boydii* (0.3%), *Shigella dysenteriae* (0.3%), *Shigella sonnei* (0.3%), *Salmonella* Typhi (0.4%) and non-typhoidal *Salmonella*. (NTS – 0.6%). *Vibrio* and *Campylobacter* were not detected. Several other organisms (*Comamonas estosterone, Brevundimonas vesicularis*, and *Pantoea agglomerans)* that had not been previously reported from clinical specimens in Kenya were identified using BD Phoenix technology. These organisms most likely represented commensal flora and not true pathogens and their identification demonstrates the potential of the BD Phoenix to identify fewer common organisms.

### Antimicrobial resistance

When the overall percentage of resistant isolates was calculated (excluding intrinsic resistance), variable resistance rates were observed among the nine antimicrobials tested (Table 2). Overall, for *Salmonella spp*. and *Shigella spp*, ampicillin had the highest resistance rate (83%) followed by tetracycline (50%) while meropenem and Amoxiclav were the least resistant, and Ciprofloxacin was 100.0% susceptible to both.

**Table 1.** Prevalence of pathogenic bacteria isolates recovered from patient’s stool samples in Kawangware informal settlement, Nairobi, Kenya, Nov 2018 to Jan 2020.

| Bacteria identified | number | % |
| --- | --- | --- |
| *Shigella flexneri* | 35 | 58.3 |
| Non-typhoidal *Salmonella* | 8 | 13.3 |
| *Salmonella ser. Typhi* | 5 | 8.3 |
| *Shigella boydii* | 4 | 6.7 |
| *Shigella sonnei* | 4 | 6.7 |
| *Shigella dysenteriae* | 4 | 6.7 |

**Table 2.0.** Antimicrobial resistance of *Salmonella* and *Shigella isolates* recovered from patients’ stool samples in Kawangware informal settlement, Nairobi. Phenotypes and Gene profiles.

|  | **Antibiotic** |  |  |  |  |  |  |  |  |  |  |  |
| --- | --- | --- | --- | --- | --- | --- | --- | --- | --- | --- | --- | --- |
| **Bacteria** | Ampicillin (%rest) | Tetracycline(%rest) | Cotrimoxazole(%rest) | Erythromycin(%rest) | Meropenem(%rest) | Chloramphenicol(%rest) | Ceftriaxone(%rest) | Gentamycin(%rest) | Ciprofloxacin(%rest) | Rifampin(%rest) | Amoxiclav(%rest) | # isolates tested |
| *S. flexneri* | 33(94) | 15(43) | 15(43) | 11(31) | 1(3) | 7(20) | 3(9) | 2(6) | 0(0) | 9(26) | 0(0) | 35 |
| *S. typhi* | 3(60) | 3(60) | 2(40) | 3(60) | 0(0) | 1(20) | 2(40) | 0(0) | 0(0) | 0(0) | 0(0) | 5 |
| NTS | 5(63) | 5(63) | 4(50) | 2(25) | 0(0) | 1(13) | 1(13) | 1(13) | 0(0) | 0(0) | 1(13) | 8 |
| *S. dysenteriae* | 3(75) | 3(75) | 2(50) | 0(0) | 0(0) | 1(25) | 0(0) | 0(0) | 0(0) | 0(0) | 0(0) | 4 |
| *S. boydii* | 3(75) | 2(50) | 3(75) | 3(75) | 0(0) | 0(0) | 0(0) | 0(0) | 0(0) | 0(0) | 0(0) | 4 |
| *S. sonnei* | 3(75) | 2(50) | 2(50) | 1(25) | 0(0) | 0(0) | 0(0) | 0(0) | 0(0) | 0(0) | 0(0) | 4 |
| **%age resistance** | **83** | **50** | **47** | **33** | **2** | **17** | **10** | **5** | **0** | **15** | **2** | **60** |

### Multidrug resistance to pathogenic Bacteria

*Salmonella* and *Shigella* isolates were resistant to multiple antimicrobials (Table 2).

### Phenotypes and Gene profiles

A total of 60 isolates representing six different phenotypes detected in the study were tested by PCR for ARG determinants. Correlation was observed between genotype and phenotype (Table 3). The most detected genes were *tet(A), tet (B), sul1* and *sul2, aac*, and ampC. The least common was *bla*_*TEM*_. We did not detect the *tet*C gene.

**Table 3.** Phenotypes and genes for different antibiotic resistant pathogenic bacteria isolates recovered from patients’ stool samples in Kawangware informal settlement, Nairobi.

| Isolate | Phenotype | Resistance genes |
| --- | --- | --- |
| *S. Typhi* | TXT, AMP, GEN | *tetA,tetB,sul1,sul2,aac(3), ampC* |
| *Salmonella spp* | TET, GEN | *tetA,tetB, aac(3), ampC* |
| *S. boydii* | TXT, TET, GEN | *tetA,tetB,sul1,sul2, aac(3), ampC* |
| *S. dysenteriae* | TXT, TET, GEN | *tetA,tetB,sul1,sul2, aac(3), ampC* |
| *S. flexneri* | TXT, TET, GEN | *tetA,tetB,sul1,sul2, aac(3) , ampC* |
| *S. sonnei* | TXT, TET, GEN | *tetA, tetB, sul1, sul, aac(3), ampC* |

*Sulfamethoxazole and trimethoprim (TXT); Tetracycline (TET); Ciprofloxacin (CIP); gentamicin (GEN); ceftriaxone (CXM); Augmentin (AUG); chloramphenicol (CLO) Ampicillin (AMC); tetracycline resistance genes (tetA, tetB); Sulfonamide resistance genes (sul1, sul2); Aminoglycoside resistance genes aac(3), ; β-lactamase resistance genes ( bla<sub>TEM</sub>, bla<sub>SHV</sub>); ampicillin resistance genes (ampC)*

### Detection of phenotypes and genes for antimicrobial resistance in bacteria isolates recovered from patients’ stool samples in Kawangware informal settlement, Nairobi

Antimicrobial resistance phenotypes can be linked to specific genetic elements with multiple different phenotypes and their related genotypes detected in this study (Table 3). The detection of the presence of phenotypes and genes for antimicrobial resistance in different bacteria indicates that antimicrobial use can be selected for bacteria with novel or certain resistance determinants. In this study; the presence of *tet(A,B)*gene and tetracycline resistance had 93.0% agreement, the presence of *(aac(3)* and gentamycin resistance 80.0%, the presence of *sul*1 and *sul*2 and cotrimoxazole resistance (91.0%), presence of *ampC* and ampicillin resistance 85.0%, and presence of *bla*_*TEM*_, *bla*_SHV_ and ceftriaxone resistance (89.0%). However, it was also observed that *S. boydii* and *S. flexneri* had resistance genes to ampicillin, though the organisms were sensitive to this antibiotic when tested by MIC. The resistance to this antibiotic was likely not expressed phenotypically or weakly expressed.

## Discussion

This study aimed to determine the prevalence and antimicrobial resistance of *Salmonella typhi*, nontyphoidal *Salmonella, Shigella, Vibrio*, and *Campylobacter* and resistance determinants present in these organisms in an urban informal settlement in Nairobi, Kenya. In this study, resistance to commonly used antibiotics such as ampicillin, co-trimoxazole, and chloramphenicol was high compared to fluoroquinolones and third-generation cephalosporins.

While the observed rates of resistance to first-line antibiotics were slightly lower compared to other studies, there was no resistance to fluoroquinolones and low resistance to cephalosporins by both *Salmonella* and *Shigella*. [4,8,24,28-34]. Despite a high prevalence of ciprofloxacin resistance in the region, our findings suggest the prevalence of ciprofloxacin resistance was low in this population, implying that in situations where antimicrobial therapy is indicated, fluoroquinolones may be effective empirical treatment in patients from this informal settlement. Moreover, the results of this study reinforce earlier studies showing a high frequency of resistance to cotrimoxazole [4,8,24,29-34]. The study identified a range of resistance determinants conferring resistance to β-lactams (bla _*TEM*_ and bla _*SHV*_), aminoglycosides (acc(3), folate pathway inhibitors (sulfonamide resistance genes) *sul*1, *sul*2 and tetracyclines *tet*(A) and *tet*(B).

In contrast to previous studies from Kenya and the Southern African region which reported high levels of AMR among Enterobacterales such as *Salmonella* Typhi, *K. pneumonia, E. coli, S. flexneri, S. dysenteriae, S. sonnei* isolated from humans, livestock, the environment and sewage [4,8,24,29-34], our study revealed reduced frequencies of antimicrobial resistance. A further description of some of the other study findings showed these isolates did not have a high frequency of resistance to fluoroquinolones, third-generation cephalosporins, and aminoglycosides that was previously reported in Kenya [8,9,34], and the high prevalence of strains resistant to more than three antibiotics (CEF, CIP, GEN, TET, TXT), our study indicated lower or reduced frequencies of resistance. Our results are consistent with Langata *et al* [32] where antimicrobial resistance among *S*. Typhi isolates in Nairobi was low. The reduced frequencies of resistance to different antimicrobials in our study could be attributed to improved sanitary practices that included the use of toilets, frequent hand washing after toilet use, and the use of piped and treated drinking water that minimizes the acquisition and spread of resistant microorganisms. Antimicrobial stewardship remains crucial. While most of the enteric *Salmonella* and *Shigella* infections in otherwise healthy individuals do not require antimicrobials, treatment can be lifesaving in severe disease or patients with co-morbidities. When treatment is indicated, these findings indicate the possibility that a wider range of commonly available antimicrobials (fluoroquinolones and 3^rd^ generation cephalosporins) may remain effective.

The results of the current study, show that the prevalence of pathogenic Enterobacterales recovered from patients in this informal settlement was similar to that reported in other informal settlements such as Kibera [25], Mukuru [34-36], other regions of the country [8] and sub-Saharan Africa [1,37,38].

There was an association between genes detected and specific resistance phenotypes of the bacteria detected in this study. *tet(A), tet(B), sul1*, and *sul2* were the most detected antimicrobial resistance genes in this study and were associated with previously described resistance phenotypes. The resistance determinants to cephalosporins and aminoglycosides corresponded to those described elsewhere [24]. In this study, the presence of the *bla*TEM gene was phynotypically associated with phenotypic resistance to ampicillin, while the carriage of *bla*SHV was associated with third-generation cephalosporins resistance. The presence of *tet(A)* and *tet(B)* resistance genes was phynotypically associated with tetracycline resistance while the presence of *sul1* and *sul2* were associated with phenotypic resistance to cotrimoxazole (*sul*3 was not detected in this study). This finding was consistent with published studies from Canada, Europe, Australia, and Korea [39-41] that showed similar phenotypic-genotypic correlations. The Australian study found that blaTEM-1-carrying bacteria were resistant to beta-lactam/beta-lactamase inhibitor combinations such as amoxicillin-clavulanate or ampicillin-sulbactam. The resistance mechanism was attributed to the TEM β-lactamase hyperproduction as reported by Waltner -Toews *et al* [24]. Detection of strains carrying blaTEM but not the *bla*TEM-1 or *bla*TEM-2 genes suggests penicillin-resistant strains may carry *bla*TEM-type ESBL derivatives. There could have been other resistance mechanisms that were present but were not tested for in this study, such as active efflux pumps which is one of the mechanisms of resistance in Enterobacterales [15]. However, two species of *Shigella* (*S. boydii* and *S. flexneri*) exhibited resistance genes for ampicillin though the isolates remained sensitive to the antibiotic when tested by disc diffusion method. This suggests the need for active surveillance and use of affordable and advanced test methods such as BD Phoenix M50 which is provided on placement terms by BD in Kenya that can detect both genotypic and phenotypic antibiotic resistance in relatively shorter time compared to conventional methods that will take a minimum of three days. This could also solf the problem possed by those bacteria that fail to express resistance phynotypically yet they habor resistance genes for such antimicrobials.

The frequency of phenotypic resistance to first-line drugs (i.e. chloramphenicol, ampicillin, tetracycline) among *Salmonellae* and *Shigella*, isolates in this study was lower than the resistance rates reported in other studies [4,34,36]. Several isolates as indicated in table 2 were resistant to gentamicin, chloramphenicol, and trimethoprim-sulfamethoxazole. The two antibiotics (ampicillin, chloramphenicol) commonly used in the empirical management [42] of diarrhea are associated with high rates of resistance among *Salmonella* and *Shigella*. Efficacy was maintained for ceftriaxone and is typically reserved for MDR cases [3,4]. Third-generation cephalosporins (ceftriaxone) and fluoroquinolones resistance was 10% and 0% respectively implying reduced resistance in contrast to studies done elsewhere [15,34,38] and supporting their use for empirical treatment when clinically indicated. However, *S flexneri* and *S. boydii* had high resistance to ampicillin, tetracycline, and cotrimoxazole. The use of third-generation cephalosporins as second-line treatment may in turn play a role in the emergence of resistance to β-lactam antibiotics [39]. The resistance to β-lactam antibiotics may be the result of a co-selection from another family of antibiotic resistance mechanisms located on the same genetic mobile element [4]. MDR in this setting could also be associated with the movement of people with resistant microorganisms which may influence the dissemination of Extended Spectrum β-Lactamases (ESBLs)-carrying isolates in the community similar to what was observed with New Delhi metallo-β-lactamase in Pakistan and extensively drug-resistant *Salmonella enterica* serovar Typhi respectively in the United Kingdom [13,43]. Historically, human migration between the Manhiça district in Mozambique and the neighboring republic of South Africa supported this dissemination or movement of resistant traits from one region to another [44]. The said population movement was confirmed to have contributed to the increase of cephalosporin resistance in the Republic of South Africa [44].

This study had a few limitations. Notably, *E coli* though not always a pathogen was the most abundant bacteria isolated in the study. However, the additional pathotyping required to fully characterize the *E. coli* strains was beyond the scope of this study. Pathotype characterization would have allowed us to determine the proportion of specimens with pathogenic *E. coli* and further understand the distribution of *E. coli* pathotypes in this study and any associated AMR phenotypes and potential medical implications in the study area. Patient recruitment was also limited in some cases by lack of consent from the head of the household.

## Conclusion

In this study, we observed low resistance frequencies for fluoroquinolones and cephalosporins compared to other studies in Kenya and some parts of Africa. This suggests the possibility that a wide range of antimicrobial agents may be available for the empirical management of infections due to these organisms. Even though these findings will inform healthcare providers on the management of bacterial infections they cannot be generalized for the entire country.

## Acknowledgment

The authors would like to acknowledge CDC-DGHP HQ for funding that enabled us to carry out this study successfully. In addition, the KEMRI-CDC Laboratory staff for their cooperation during the study and Riruta Satellite Hospital management and staff for allowing us to conduct this study in their institution.

## Disclaimer

The findings and conclusions in this report are those of the authors and do not necessarily represent the official position of the United States Centers for Disease Control and Prevention.

## Author contributions

**Conceptualization:** Bonventure Juma, Matthew Mikoleit, Elizabeth Hunsperger, Newton Wamola,Samuel Kariuki

**Data curation:** Newton Wamola, Isaac Owuor, Michael Osita

**Formal analysis:** Bonventure Juma, Newton Wamola

**Funding acquisition:** Elizabeth Hunsperger, Bonventure Juma

**Investigation:** Newton Wamola, Michael Osita, Caroline Apondi

**Methodology:** Bonventure Juma, Matthew Mikoleit, Newton Wamola,, Caroline Apondi

**Project administration:** Bonventure Juma, Newton Wamola, Michael Osita,

**Resources:** Bonventure Juma, Elizabeth Hunsperger

**Supervision:** Bonventure Juma, Newton Wamola

**Validation:** Bonventure Juma, Newton Wamola, Michael Osita,

**Writing original draft:** Bonventure Juma, Michael Osita, Newton Wamola

**Writing review and editing** Elizabeth Hunsperger, Mathew Mikoleit, Bonventure Juma, Newton Wamola,Samuel Kariuki

## Supporting information

We have included the map of Darogetti Division.

